# The impact of goggle-associated harms to health and working status of nurses during management of COVID-19

**DOI:** 10.1101/2020.05.11.20094854

**Authors:** He Xiao-huan, Feng Yan-ru, Li Gao-ming, Pang Xiao-jiao, Chen Ting, Zhou Ya-li, Zhang Hao, Lang Jing, Li Li-min, Feng Li, He Xin, Zheng Wei, Miao Hong-ming, Wang Yong-hua, Kang Xia

**Author notes:** He Xiao-huan, Feng Yan-ru, Li Gao-ming, Pang Xiao-jiao contributed equally to this article. Email address: He Xiao-huan,; Feng Yan-ru,; Li Gao-ming,; Pang Xiao-jiao,; Chen Ting,; Zhou Ya-li,; Zhang Hao,; Lang Jing,; Li Li-min,; Feng Li,; He Xin,; Zheng Wei,; Miao Hong-ming,; Wang Yong-hua,; Kang Xia. **Correspondent authors: Kang Xia^1^, Wang Yong-hua^1^, Miao Hong-ming^3^ and Zheng Wei^1^:** 1. Department of Orthopedics, General Hospital of Western Theater Command, Rongdu Avenue No. 270, Chengdu, 610000, PRC; 3. Army Medical University.

## Abstract

**Background:** To investigate the impact of goggles on their health and clinical practice during management of patients with COVID-19.

**Methods:** 231 nurse practitioners were enrolled who worked in isolation region in designated hospitals to admit patients with COVID-19 in China. Demographic data, goggle-associated symptoms and underlying reasons, incidence of medical errors or exposures, the effects of fog in goggles on practice were all collected. Data were stratified and analyzed by age or working experience. Risk factors of goggle-associated medical errors were analyzed by multivariable logistical regression analysis.

**Findings:** Goggle-associated symptoms and foggy goggles widely presented in nurses. The most common symptoms were headache, skin pressure injury and dizziness. Headache, vomit and nausea were significantly fewer reported in nurses with longer working experience while rash occurred higher in this group. The underlying reasons included tightness of goggles, unsuitable design and uncomfortable materials. The working status of nurses with more working experience was less impacted by goggles. 11.3% nurses occurred medical exposures in clinical practice while 19.5% nurses made medical errors on patients. The risk factors for medical errors were time interval before adapting to goggle-associated discomforts, adjusting goggles and headache.

**Interpretation:** Goggle-associated symptoms and fog can highly impact the working status and contribute to medical errors during management of COVID-19. Increased the experience with working in PPE through adequate training and psychological education may benefit for relieving some symptoms and improving working status. Improvement of goggle design during productive process was strongly suggested to reduce incidence of discomforts and medical errors.

## Introduction

Over 2.4 million people have been diagnosed with coronavirus disease 2019 (COVID-19) till now. With the global outbreak of COVID-19 in recent three months, medial resources, including personal protective equipment (PPE) and human resources, are insufficient to cope with rapid growth of diagnosed patients. Recently, the health status of healthcare professionals involved in management of patients with COVID-19 was concerned^[1-5]^. A number of physicians realized and called on the potential risks on health of healthcare professionals due to the shortage of PPE^[4-9]^. On the other hand, donning of the PPE often associates with uncomfortable and cumbersome. It was reported that PPE can cause headache and difficulty breathing, indicating PPE itself may potentially harm to healthcare professionals in some situations^[10]^, thus PPE-associated harms can not be neglected during COVID-19 pandemic.

A special situation that should be considered is many healthcare professionals whose specialties are not respiratory or infective diseases involve in the management of patients with COVID-19 due to the sudden outbreak of COVID-19 worldwide. Under the emergent situation, a number of them got limited training on wearing PPE and had little time to adapt to working in PPE before going to frontline. It is reasonable that insufficient experience with working in PPE combined with the states of fearing exposure may enhance the discomfortableness. These discomforts may negatively impact the working status and the accuracy of procedures. However, whether and to what extent the negative effects of PPE on both healthcare professionals and patients were rarely reported in studies on COVID-19.

Comparing with physicians, nurses have more chances to directly contacts with patients in daily practice. The possibility of occurring medical exposures during clinical practice may be increased. Although nurses who involved in management of this pandemic received training on use of PPE and psychological education before they went to frontline^[11]^, high proportion of nurses that engaged in the management of epidemic still complained about discomforts associated with PPE, of which goggles were mostly mentioned in all kinds of PPE. Nurses indicated that goggles may cause serious somatic symptoms that remarkedly impacted their working status. Meanwhile, fogging in goggles was likely unavoidable during clinical work and their vision was interfered which impacted their clinical practice. Considering the universality and necessities of wearing goggles during this pandemic, it is essential to investigate the epidemiologic features of goggle-associated discomforts and to what extent the goggles impact the working status of nurses. In this questionnaire-based retrospective study, we analyzed 231 questionnaires getting from nurse practitioners. All of participants worked in isolation region for management of patients with COVID-19. We detailly interpreted the prevalence and distribution of goggle-associated complications and interpreted the underlying reasons through stratified age or working experience. Furthermore, the risk factors associated with medical errors were analyzed. On the other hand, the negative effects of fog in goggles on working status of nurses were studied. We still introduced some improving measures that can reduce the goggle-associated discomforts. Our findings specifically emphases the health problems caused by goggles during management of patients with COVID-19. In addition, we would like to arouse the concerns from authorities and PPE manufactures on the harms caused by PPE to both healthcare professionals and patients during the management of COVID-19.

## Methods

### Study design and participants

This study was approved by the Ethical Committee at General hospital of Western Theater Command. This questionnaire-based retrospective study was done at several designated hospitals to treat patients with COVID-19 in Wuhan and Chongqing, China. All participants were nurse practitioners, who worked in isolation region (also called red zone) from late January to late March and participated in the direct management of patients with COVID-19. The survey was performed anonymously, the data collected were presented in an aggregated form.

### Definitions and data collection

Medical errors in this study are defined as unsuccessful clinical procedures, such as unsuccessful venipuncture, or medication errors on patients and harms to healthcare professionals. Medical exposure specifically indicated the situation that healthcare professionals were exposed to the environment that could be infected due to some unexpected reasons, such as needle-stick injuries, damage of PPE, etc. Skin medical device-related pressure injury SMDRPI means localized skin injury as a result of sustained pressure from medical device, it can be divided into 4 stages as defined in previous study: Stage 1, whole skin with erythema that does not whiten; Stage 2, loss of skin in its partial thickness with dermis exposure; Stage 3, loss of skin in its overall thickness; Stage 4, loss of skin in its total thickness and tissue loss^[12]^.

Data were collected and evaluated using own-designed questionnaires. Any missing or uncertain information was excluded during data process. We primarily concentrated on two aspects: discomforts caused by goggles and the effects of fog in goggles on clinical practice. Considering ages and practice experience may be two critical impact factors to influence metal stress, working status and tolerance for symptoms, critical information was further analyzed stratified by age groups and practice durations.

Data on age, sex, specialty, duration of practice, experience of wearing goggles, mental stress in clinical practice, goggles associated symptoms (claustrophobia, irritability, vomit, headache, dizziness, nausea, eczema, skin pressure injury), effects of fog in goggles on medical exposure, measures relieving discomfort and fog, were collected and evaluated.

### Statistical analysis

Continuous variables were expressed as median and interquartile range (IQR), comparisons between two groups or multiple groups were done with Mann-whitney *U* or kruskal-wallis *H* test, respectively. Categorical variables were presented as number (%) and differences among groups were compared with Chi-square or Fisher’s exact test where appropriate. The relationships between level of tightness of goggles and level of mental stress and between age and practice duration were assessed by Spearman’s rank correlation coefficient, respectively. The potential factors associated with medical errors were investigated using univariate and multivariate binary logistic regression models. Statistical analyses in this study were down with SPSS, version 18.0 (SPSS statistics for windows, Inc., Chicago, Ill., USA). All tests were bilateral and p values less than 0.05 were considered significant.

## Results

### Most nurses had insufficient experience to work in PPE

Total 231 questionnaires were eligible for subsequent analysis. The median age was 32.0 (IQR 28.0-37.0) years old. The median practice duration was 11.0 (IQR 6.5-16.0) years. 96.1% (222) participants were female, the ratio of male and female showed no significance when stratified by age range or practice duration (Table 1). The specialties of participants included neurology, outpatient department, infectious disease, orthopedics, oncology, emergency, gastrointestinal surgery, respiratory, intensive critical care, ear-nose-throat (ENT), anesthesiology, obstetrics and gynecology (OG), pediatrics, minimally invasive surgery, rehabilitation, geriatrics, nephrology, cardiology, operating room, endocrine, thoracic surgery, urinary surgery, physiotherapy, burns, maxillofacial surgery and hepatology, approximately 9.1% of participants worked in department of infectious diseases or department of respirator before this pandemic. Meanwhile, only 10.0% (23) participants had the experience of wearing PPE in clinical practice and there was no significance among each group stratified by age or practice duration (Table 1). All of participants received training for wearing PPE and psychological education before going to frontline, the training hours were similar among each group, the median time for training and education in total participants was 7.0 (IQR 2.0-10.0) days (Table 1). All of participants worked for average 4 (IQR 4 - 5) hours in isolation region per day. To evaluate the level of mental stress on fearing exposure, participants were asked to select a point from an “1-10” scale (1 = Not at all, 10 = Extremely), the median point was 5.0 (IQR 3.0-8.0). Comparing with other two age groups, this rating was significantly lower in nurses with age ≥ 40 years (4.0, 1.0-6.5, p = 0.032) (Table 1).

**Table 1:** Demographic and baseline characterizes of enrolled participants.

|  | Total<br>(n=231) | Age range, years |  |  |  | Practice duration, years |  |  |  |
| --- | --- | --- | --- | --- | --- | --- | --- | --- | --- |
|  |  | 20-29<br>(n=73) | 30-39<br>(n=129) | ≥40<br>(n=29) | p value | 0-10<br>(n=112) | 11-20<br>(n=100) | ≥ 21<br>(n=19) | p value |
| Sex |  |  |  |  |  |  |  |  |  |
| male | 9 (3.9%) | 4 (5.5%) | 4 (3.1%) | 1 (3.4%) | 0.709 | 8 (7.1%) | 1 (1.0%) | 0 (0%) | 0.073 |
| female | 222 (96.1%) | 69 (94.5%) | 125 (96.9%) | 28 (96.6%) |  | 104 (92.9%) | 99 (99.0%) | 19 (100%) |  |
| Duration in isolation<br>region per day, hours | 4 (4-5) | 4 (4-6) | 4 (4-4) | 4 (4-6) | 0.176 | 4 (4-6) | 4 (4-4) | 4 (4-6) | 0.002 |
| Experience of<br>wearing goggles | 23 (10.0%) | 7 (9.6%) | 12 (9.3%) | 4 (13.8%) | 0.760 | 12 (10.7%) | 9 (9.0%) | 2 (10.5%) | 0.914 |
| Training time of<br>wearing goggles, days | 7 (2-10) | 7 (1-10) | 7 (2-10) | 5 (1-9) | 0.507 | 7 (1-10) | 7 (3-10) | 5 (1-7) | 0.180 |
| Mental stress on<br>fearing exposure | 5.0 (3.0-7.0) | 5.0 (3.0-7.0) | 5.0 (3.5-8.0) | 4.0 (1.0-6.5) | 0.032 | 5.0 (3.0-7.0) | 5.5 (3.0-8.0) | 4.0 (1.0-6.0) | 0.173 |
| Data are median (IQR) or n (%). p values were calculated by kruskal-wallis $H$ test, $\chi^2$ test, or Fisher’s exact test, as appropriate. | | | | | | | | | |

These baseline characteristics indicated most of nurses who involved in first line of management of COVID-19 had little experience in working in PPE before this pandemic. Due to the urgent situation, there was limited time for training on wearing PPE and receiving psychological education.

### Goggle-associated symptoms were highly prevalent in nurses who involving in management of COVID-19

To investigate to what extent goggles impacted nurses, we analyzed the symptoms caused by wearing goggles that were reported by nurses. The occurred symptoms, the initial symptoms and the most frequently occurred symptoms were collected and analyzed, respectively.

In our report, the three most reported occurred symptoms were headache (79.1%), skin pressure injury (I – IV stages) (66.2%) and dizziness (49.4%), all of symptoms showed no significant differences among age groups (Table 2). When stratified by practice duration, the incidence of headache and vomit was similar between nurses with 0-10 years working experience and 11-20 years working experience, but much higher than the incidence in group with ≥21 years working experience. On the contrary, the incidence of rash was highest in nurses with ≥ 21 years working experience comparing with other two groups (Table 2). On the other hand, headache, skin pressure injury, nausea and dizziness were the symptoms with highest incidence in each age group or practice duration group (Fig 1a and 1d).

**Table 2:**
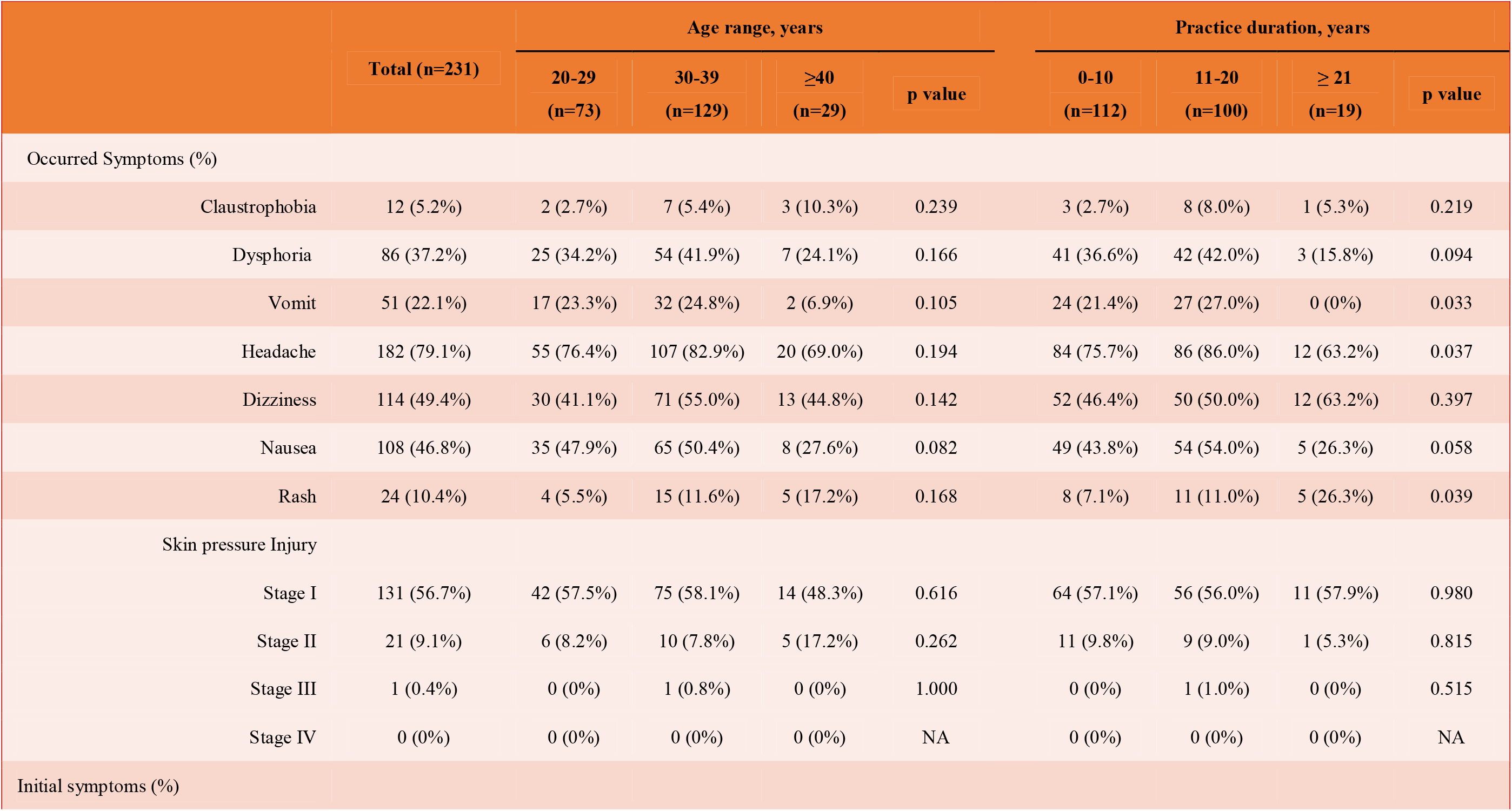
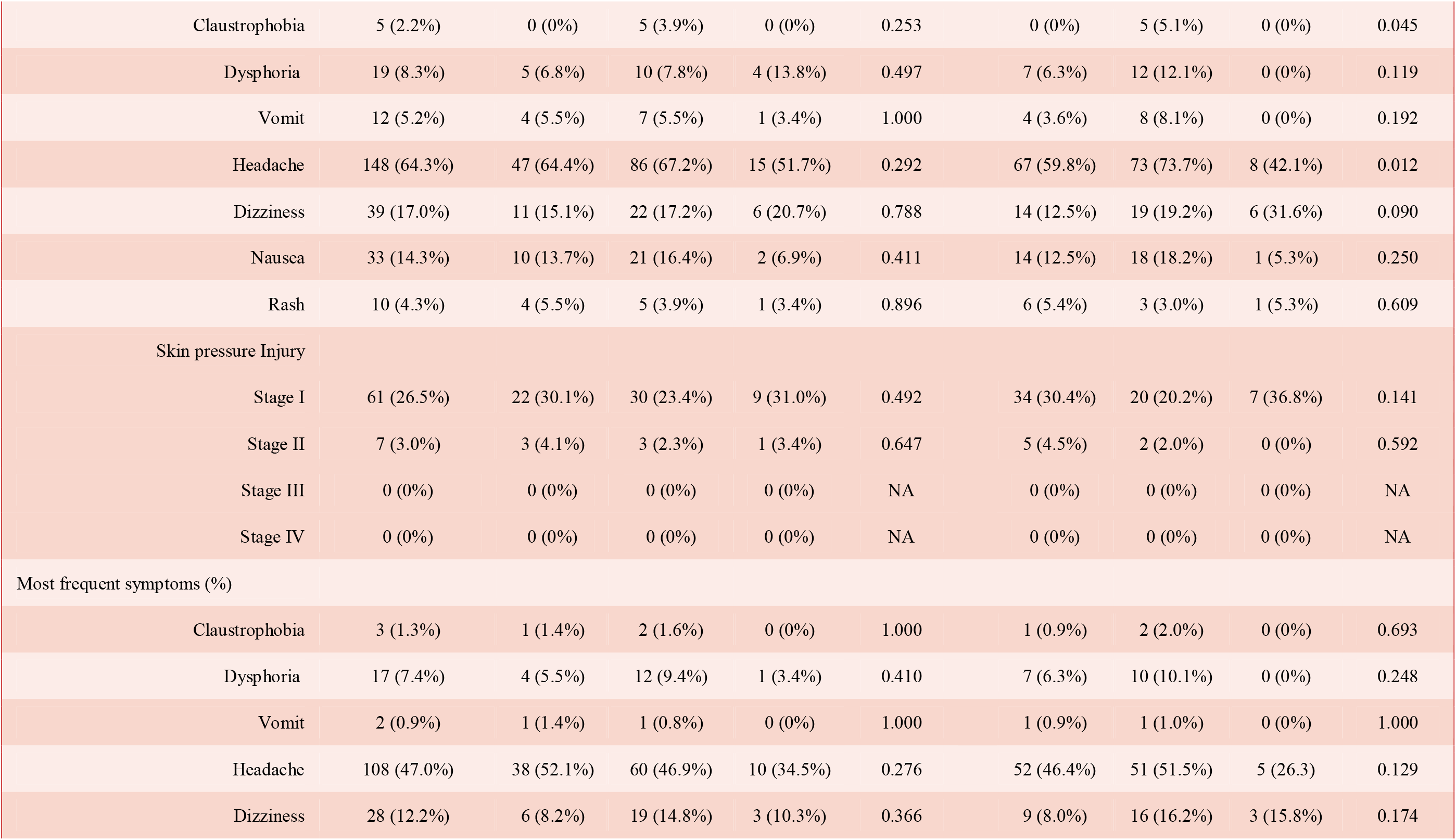
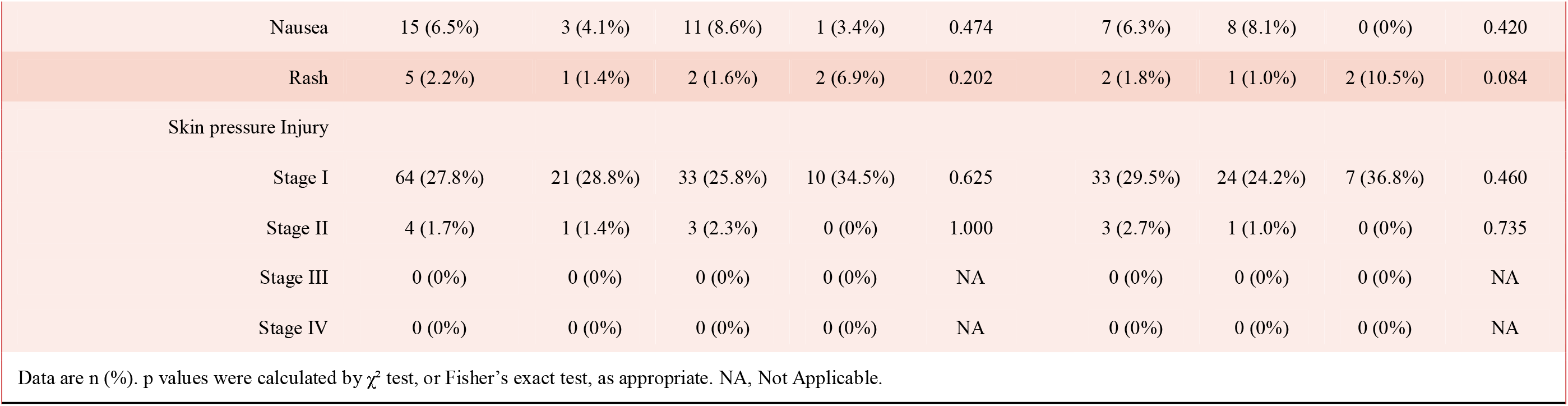
Goggle-associated symptoms.

**Figure 1.**
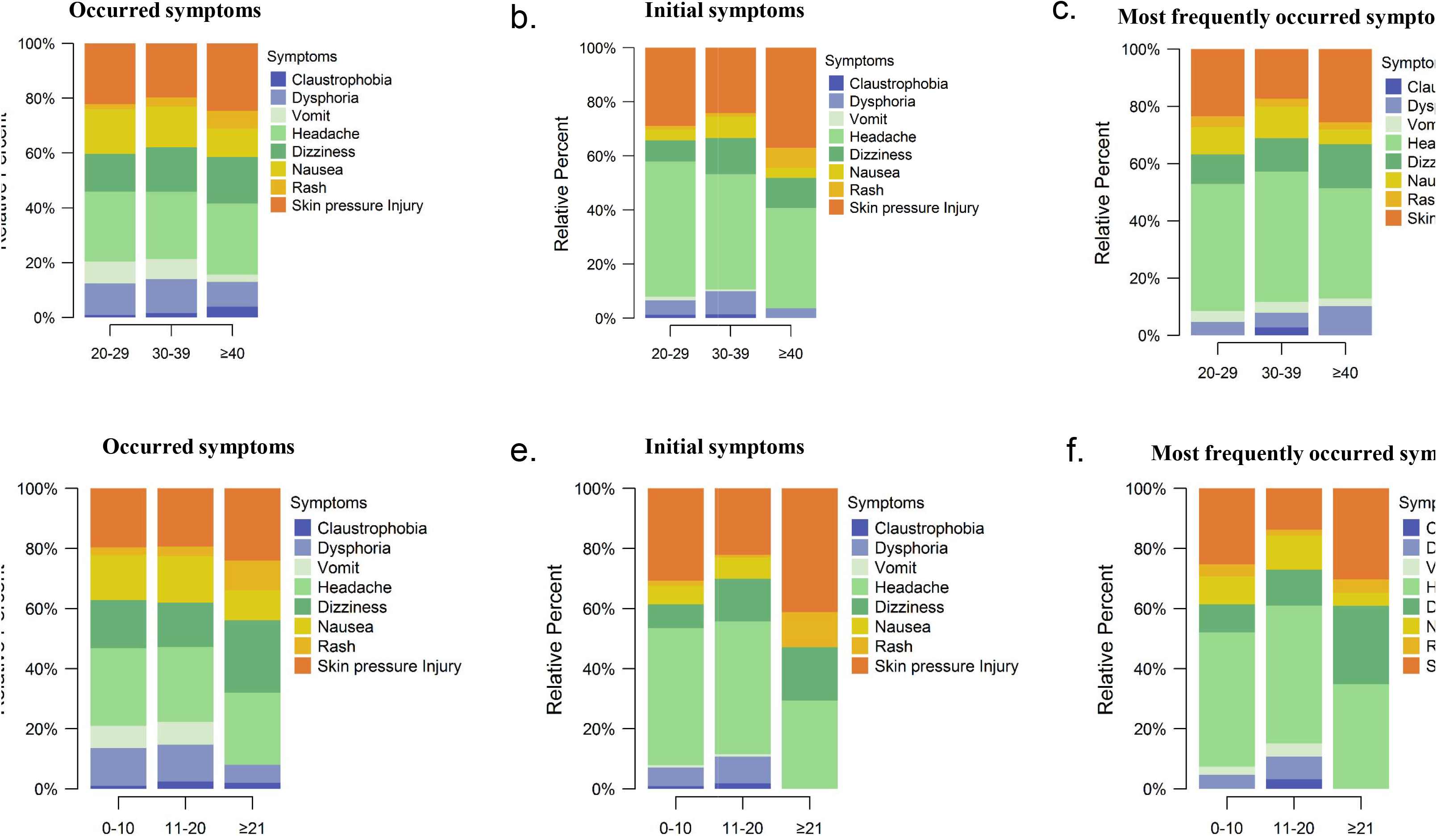

When asked the earliest occurred symptoms, headache was also mostly reported (64.3%) and the proportion was much higher than other symptoms. In addition, the proportion of initial symptom as headache in nurses with ≥21 years working experience was much lower than other two groups (42.1%, p=0.012). The second mostly reported initial symptom was skin pressure injury (I - IV stages) and the third one was dizziness. Besides to headache and claustrophobia, no significance was observed in each symptom when stratified by age or practice duration (Table 2). When analyzing the distribution of each symptoms in age group or practice duration group, the percentages of headache and skin pressure injury were much higher than other symptoms (Fig 1b and 1e).

Next, we further investigated the most frequent occurred symptoms. Headache was also reported as the symptom on rank 1 that was most frequently occurred (47.0%). The second was skin pressure injury and the third symptom was dizziness. In addition, the proportion of headache was nearly 20% higher than the proportion of skin pressure injury. Furthermore, the proportion of all symptoms showed no significant differences in each group (Table 2). Similar with the distributions in occurred symptoms and initial symptoms, headache and skin pressure injury were the top 2 symptoms with highest proportions in each age group and practice duration group (Fig 1c and 1f).

Taking together, these results showed goggle-associated discomforts extensively presented in nurses with a high proportion. Notably, headache and skin pressure injury should be noticed with special caution while rash, vomit and nausea were also common symptoms in different groups. Moreover, vomit and headache were likely to occur in nurses with shorter working experience and rash was more likely to occur in nurses with longer working experience, indicating precautions for goggle-associated symptoms should be prepared differentially.

### Goggle-associated symptoms negatively impacted the working status

Our results showed the onset of goggle-associated discomforts occurred mostly in the first day after wearing goggles. In addition, only wearing goggles for average 3.0 (IQR 2.0-10.75) days, the nurses can begin to endure the goggle-associated discomforts (Table 3). These results showed no obvious differences among each age or practice duration group. Nevertheless, 19.6% (44) participants never adapted to these discomforts (Data not shown).

**Table 3:**
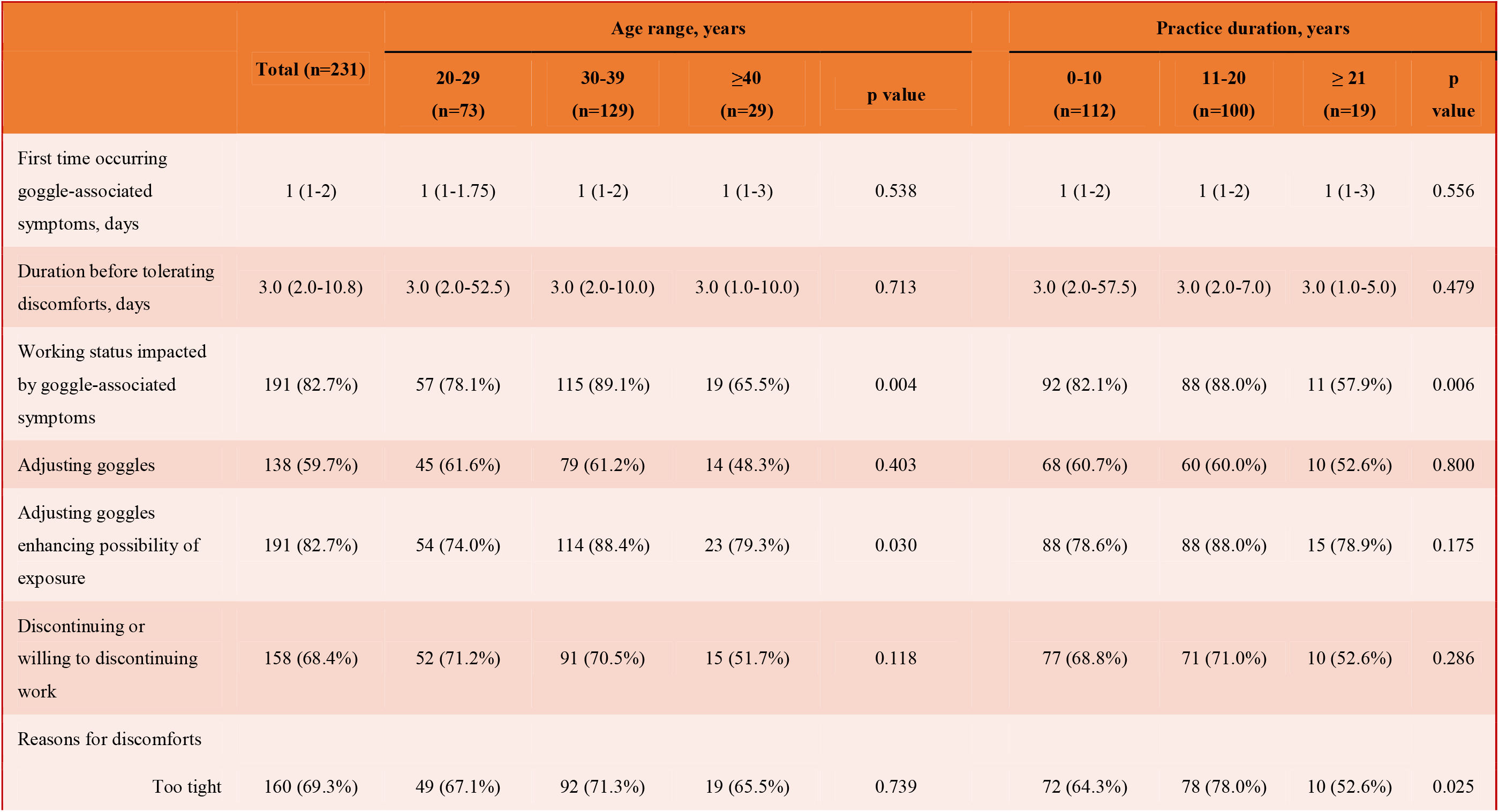
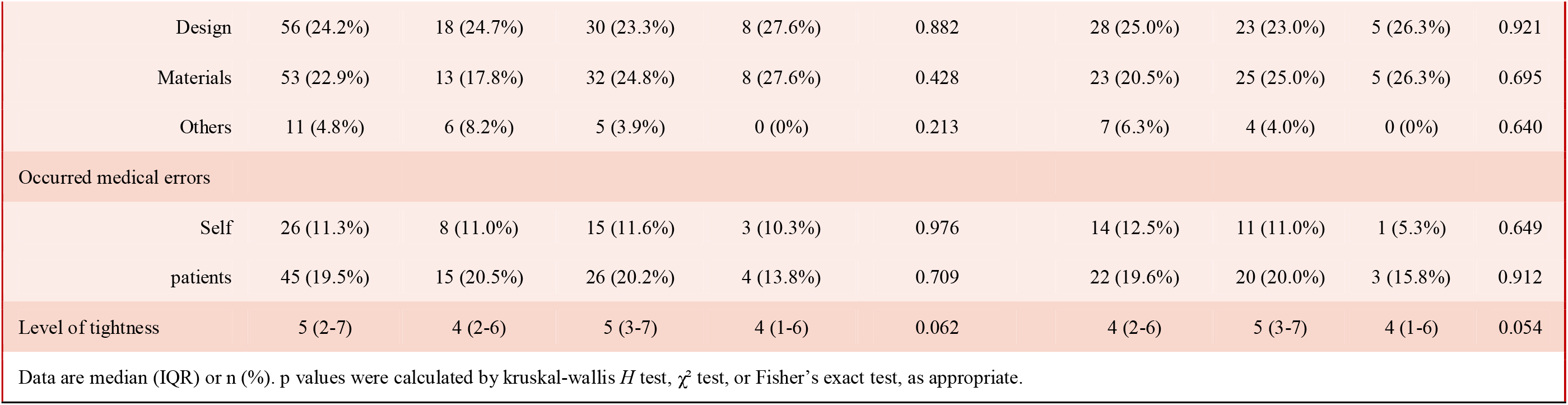
Association between goggle-associated symptoms and the working status of clinical practice.

|  | Age range, years |  |  |  |  | Practice duration, years |  |  |  |
| --- | --- | --- | --- | --- | --- | --- | --- | --- | --- |
|  | Total (n=231) | 20-29<br>(n=73) | 30-39<br>(n=129) | ≥40<br>(n=29) | p value | 0-10<br>(n=112) | 11-20<br>(n=100) | ≥ 21<br>(n=19) | p value |
| First time occurring goggle-associated symptoms, days | 1 (1-2) | 1 (1-1.75) | 1 (1-2) | 1 (1-3) | 0.538 | 1 (1-2) | 1 (1-2) | 1 (1-3) | 0.556 |
| Duration before tolerating discomforts, days | 3.0 (2.0-10.8) | 3.0 (2.0-52.5) | 3.0 (2.0-10.0) | 3.0 (1.0-10.0) | 0.713 | 3.0 (2.0-57.5) | 3.0 (2.0-7.0) | 3.0 (1.0-5.0) | 0.479 |
| Working status impacted by goggle-associated symptoms | 191 (82.7%) | 57 (78.1%) | 115 (89.1%) | 19 (65.5%) | 0.004 | 92 (82.1%) | 88 (88.0%) | 11 (57.9%) | 0.006 |
| Adjusting goggles | 138 (59.7%) | 45 (61.6%) | 79 (61.2%) | 14 (48.3%) | 0.403 | 68 (60.7%) | 60 (60.0%) | 10 (52.6%) | 0.800 |
| Adjusting goggles enhancing possibility of exposure | 191 (82.7%) | 54 (74.0%) | 114 (88.4%) | 23 (79.3%) | 0.030 | 88 (78.6%) | 88 (88.0%) | 15 (78.9%) | 0.175 |
| Discontinuing or willing to discontinuing work | 158 (68.4%) | 52 (71.2%) | 91 (70.5%) | 15 (51.7%) | 0.118 | 77 (68.8%) | 71 (71.0%) | 10 (52.6%) | 0.286 |
| Reasons for discomforts |  |  |  |  |  |  |  |  |  |
| Too tight | 160 (69.3%) | 49 (67.1%) | 92 (71.3%) | 19 (65.5%) | 0.739 | 72 (64.3%) | 78 (78.0%) | 10 (52.6%) | 0.025 |

|  |  |  |  |  |  |  |  |  |  |  |
| --- | --- | --- | --- | --- | --- | --- | --- | --- | --- | --- |
|  | Design | 56 (24.2%) | 18 (24.7%) | 30 (23.3%) | 8 (27.6%) | 0.882 | 28 (25.0%) | 23 (23.0%) | 5 (26.3%) | 0.921 |
|  | Materials | 53 (22.9%) | 13 (17.8%) | 32 (24.8%) | 8 (27.6%) | 0.428 | 23 (20.5%) | 25 (25.0%) | 5 (26.3%) | 0.695 |
|  | Others | 11 (4.8%) | 6 (8.2%) | 5 (3.9%) | 0 (0%) | 0.213 | 7 (6.3%) | 4 (4.0%) | 0 (0%) | 0.640 |
| Occurred medical errors |  |  |  |  |  |  |  |  |  |  |
|  | Self | 26 (11.3%) | 8 (11.0%) | 15 (11.6%) | 3 (10.3%) | 0.976 | 14 (12.5%) | 11 (11.0%) | 1 (5.3%) | 0.649 |
|  | patients | 45 (19.5%) | 15 (20.5%) | 26 (20.2%) | 4 (13.8%) | 0.709 | 22 (19.6%) | 20 (20.0%) | 3 (15.8%) | 0.912 |
| Level of tightness |  | 5 (2-7) | 4 (2-6) | 5 (3-7) | 4 (1-6) | 0.062 | 4 (2-6) | 5 (3-7) | 4 (1-6) | 0.054 |
| Data are median (IQR) or n (%). p values were calculated by kruskal-wallis $H$ test, $\chi^2$ test, or Fisher's exact test, as appropriate. | | | | | | | | | | |

Considering the rapid onset of discomforts and the relative long time before tolerance, we concerned whether it would impact the working status in clinical practice. As expected, total 82.7% (191) participants thought goggle-associated discomforts remarkedly impacted the working status, of which 89.1% (115) participants in 30-39 years old group thought their work was negatively influenced by goggle-associated symptoms while only 65.5% (19) participants in group of age≥40 years agreed with this viewpoint (p=0.004). Interestingly, nurses with ≥20 years working experience (57.9%) thought they were less impacted by these discomforts comparing with other two groups (p=0.006). Notably, although 82.7% (191) nurses acknowledged adjusting goggles can increase the possibility of medical exposure, 59.7% (138) nurses still adjusted goggles during working. Interestingly, among the three age groups, the proportion that acknowledging goggle-associated discomforts would increase medical exposure was lowest in nurses with age≥40 years (p=0.030). Furthermore, 68.4% (158) nurses discontinued or wanted to discontinued work due to goggle-associated discomforts and this proportion showed no significant differences when stratified by age or practice duration (Table 3). Moreover, the total times for discontinuing work were 202 (range from 1 to 10), 4.3% (10) participants discontinued working for more than 5 times.

We further explored the underlying reasons that causing goggle-associated symptoms. The feedback showed there were three primary reasons. Tightness of goggles (69.3%) was the most important reason that caused goggle-associated symptoms. Meanwhile, unsuitable design for Chinese face shape (24.2%) and uncomfortable materials, such as too stiff (22.9%) were the second reason and the third reason, respectively (Table 3).

Since it was reasonable that mental stress may make nurses wearing the goggles too tight, especially in the situation that most nurses had insufficient experience with working in PPE as well as managing infectious disease, we next explored how mental stress influenced on wearing the goggles too tight. The correlation between the two scales, “Mental stress on fearing exposure” (5.0, [IQR 3.0-7.0]) and “level of tightness” (5.0, [IQR 2.0-7.0]) was analyzed by using Spearman correlation analysis. The result showed these two indicators were moderate correlation (r = 0.651, p < 0.001) (Supple. Fig 1), indicating mental stress at least partly influenced the tightness when wearing goggles during the management of patients with COVID-19.

### Risk factors for discomfort-associated medical errors

It is reasonable that working status was likely to be influenced by discomforts and inferior working status may associate with increased possibility of medical errors, thus we collected the proportion of nurses that occurred medical errors, total 11.3% (26) nurses occurred medical exposures in clinical practice. 19.5% (45) nurses made medial errors on patients. The incidence showed no significant differences between each group when stratified by age or working experience (Table 3).

We next explored the risk factors for medical errors in management of COVID-19. Multivariable logistical regression analysis showed “Days before enduring goggle-associated discomforts” (OR, 2.73; 95% CI, 1.43-5.22), “Adjusting goggles” (OR, 2.56; 95% CI, 1.28-5.14) and “Headache” (OR, 5.50; 95% CI, 1.82-16.58) were identified as three risk factors contributing to medical errors, of which the possibility of occurring medical errors in nurses occurred headache enhanced 5.5 folders comparing with nurses without headache (Table 4).

**Table 4:** Risk factors associated with medical errors.

| Variable | Univariable OR (95% CI) | p value | Multivariable OR (95% CI) | p value |
| --- | --- | --- | --- | --- |
| Age range |  |  |  |  |
| 20-29 | 1 (ref) |  |  |  |
| 30-39 | 1.14 (0.59-2.20) | 0.701 |  |  |
| ≥ 40 | 0.80 (0.28-2.26) | 0.670 |  |  |
| Practice duration |  |  |  |  |
| 0-10 | 1 (ref) |  |  |  |
| 11-20 | 1.01 (0.54-1.86) | 0.986 |  |  |
| ≥ 21 | 0.76 (0.23-2.49) | 0.654 |  |  |
| Practice (≥ 10 vs < 10) | 1.12 (0.62-2.05) | 0.703 |  |  |
| Hours in isolation region (≥ 5 vs < 5) | 1.08 (0.56-2.07) | 0.826 |  |  |
| Experience with wearing goggles before (Yes vs No) | 1.31 (0.51-3.37) | 0.572 |  |  |
| Training time (≥ 5 vs < 5) | 1.05 (0.57-1.95) | 0.869 |  |  |
| Days before adapting to goggles (≥ 5 vs < 5) | 2.45 (1.33-4.53) | 0.004 | 2.73 (1.43-5.22) | 0.002 |
| Impact on working status by discomforts (Yes vs No) | 3.64 (1.24-10.71) | 0.019 |  |  |
| Adjusting goggles (Yes vs No) | 2.18 (1.14-4.16) | 0.019 | 2.56 (1.28-5.14) | 0.008 |
| Taking measures for discomforts | 0.92 (0.42-2.03) | 0.834 |  |  |
| (Yes vs No) |  |  |  |  |
| Claustrophobia (Yes vs No) | 2.18 (0.67-7.16) | 0.198 |  |  |
| Dysphoria (Yes vs No) | 1.62 (0.89-2.95) | 0.118 |  |  |
| Vomit (Yes vs No) | 2.08 (1.07-4.05) | 0.032 |  |  |
| Headache (Yes vs No) | 4.64 (1.59-13.55) | 0.005 | 5.50 (1.82-16.58) | 0.002 |
| Dizziness (Yes vs No) | 1.72 (0.94-3.13) | 0.077 |  |  |
| Nausea (Yes vs No) | 1.04 (0.57-1.88) | 0.900 |  |  |
| Skin press injury (Yes vs No) | 0.82 (0.45-1.51) | 0.526 |  |  |
| Rash (Yes vs No) | 0.24 (0.05-1.05) | 0.058 |  |  |
| OR, odds ratio; CI, confidence interval. ref, reference. |  |  |  |  |
| **Table 4: Risk factors associated with medical errors** |  |  |  |  |

These findings demonstrated that negative effects of goggles did not only impact the healthcare professionals’ physical and mental health, but also directly impact the practice and increase the exposure risk as well as harms to patients. In addition, the time to tolerate the discomforts, adjusting goggles and headache significantly increased the risk occurring medical errors, which should be prevented before and during management of patients with COVID-19.

### Fog in goggles may contribute to medical exposures during the management of COVID-19

Foggy goggles were reported frequently in our survey, 97.0% (224) participants met this problem during their clinical practice in management of patients with COVID-19. The median time to occurring fog in goggles was 1.0 (IQR 0.5-3.0) hours after wearing PPE. It is worthy to notice that 96.1% (222) nurses in this survey considered fog in goggles remarkedly impacted their clinical practice, however, this proportion in nurses with ≥21 years practice experience was relatively lower comparing with other groups. In addition, the average rating on the level of fog impacting the accuracy of clinical procedures was 6 (IQR 4-8), indicating most nurses though fog remarkedly impacted their working (Table 5).

**Table 5:**
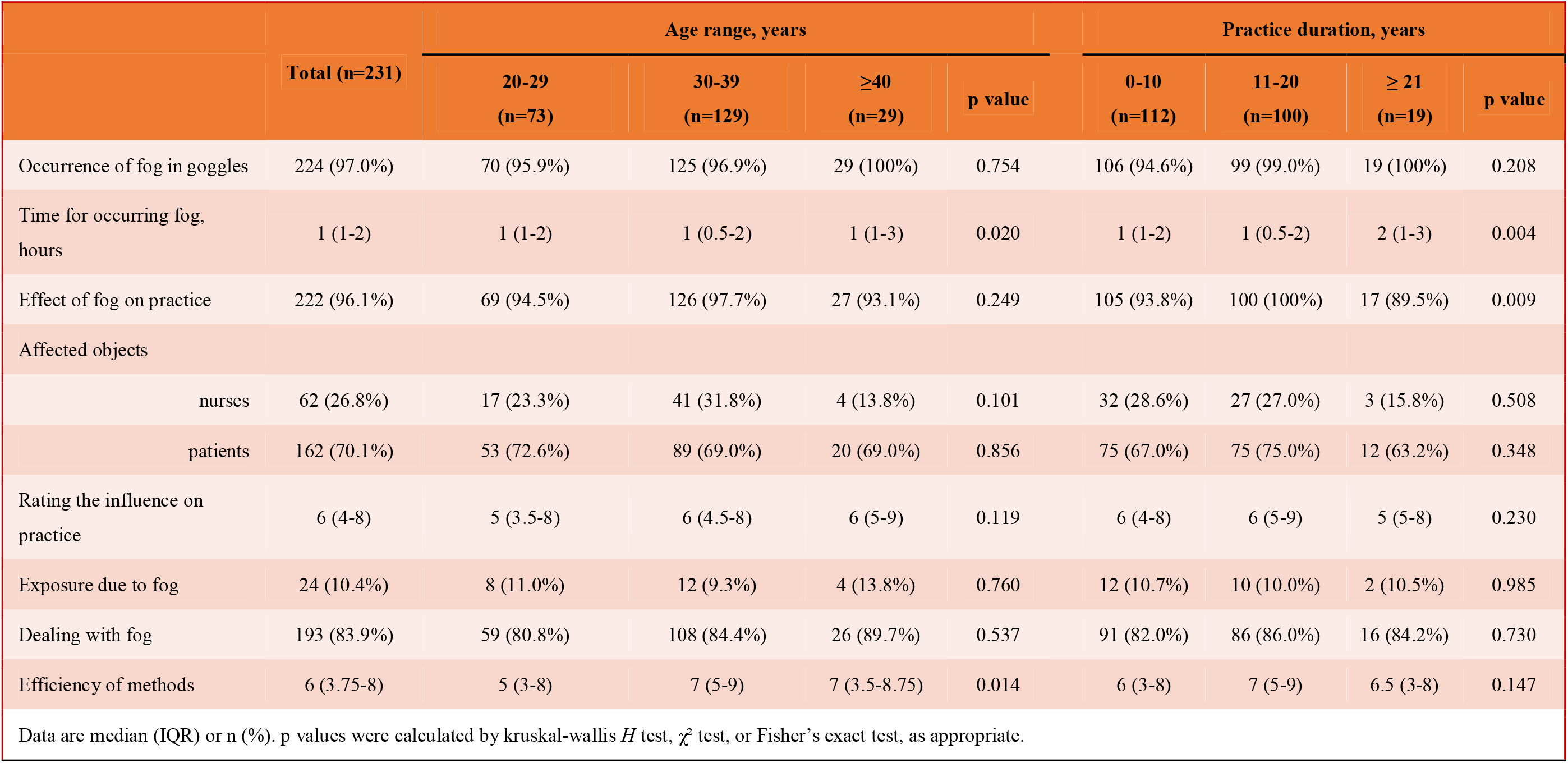
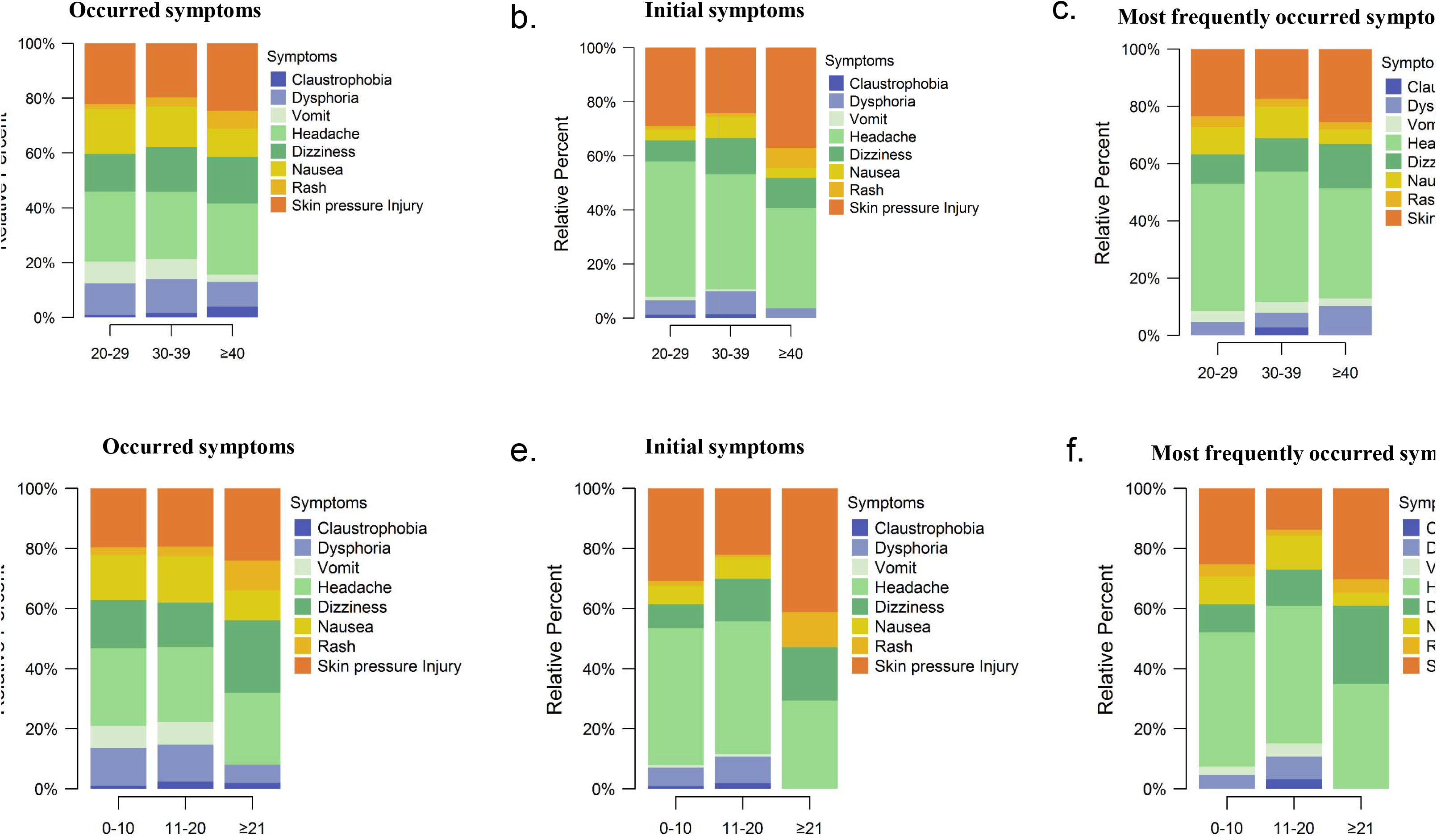
The impact of fog in goggles on clinical practice.

Interfered vision during practice caused by fog may lead to clinical exposure or medical errors, when we asked healthcare professionals or patients were more likely to be affected due to vision interfering, most participants tented to selecting patients but not healthcare professionals, and this selection showed no statistic differences among all groups when stratified by age or practice experience. Notably, 10.4% (24) participants reported they exposed during clinical practice due to fog (Table 5).

These results indicating that fog frequently occurred in clinical practice when managing patients with COVID-19. In addition, it was widely accepted by nurses in all groups that inferior vision caused by fog largely impacted the routine work and contributed to noticeable medical exposure.

### Tips for attenuating goggle-associated discomforts and relieving fog during wearing PPE

Total 83.8% (193) participants took actions to deal with fogs and this ratio showed no significant differences when stratified in age or practice experience. Varieties of reagents were utilized to smear on eyeglass, including iodine, liquid soap or soap, antifoggant, hydrogel, bath foam (Supplement Fig 3). Self-rating on the efficiency of these methods was 6 (IQR 3.75-8), interestingly, the rating in nurses under 30 years was lowest in all groups (Table 5).

There were few ways to relieve goggle-associated discomforts, most nurses directly loosed band a bit intermittently. Other methods included selecting goggles with wider or softer bands and using anti-stress trips to relieve the pressure of bands on skin, however, exposure possibility should be concerned when using these methods.

## Discussion

Due to high infectivity of SARS-CoV-2 virus, precautions on protection of healthcare professionals were frequently mentioned recently^[13,14]^. In China, best efforts were performed to prevent possible medical errors and relieve mental stress. Nurses who involved in first line treatment of patients with COVID-19 received education and training on use of PPE and personal emotion management in some hospitals^[11]^. Meanwhile, the daily working time was restricted to avoid PPE-associated complications, such as dehydration, as described by us and the other study^[11]^. However, as described in this study, due to shortage of nurses in this pandemic, the proportion of nurses who had no experience to work in PPE before this pandemic in frontline of treating COVID-19 was higher than expected. In addition, we also identified most of them did not work in department of respiratory or infective disease, implying they may have insufficient experience on fulminating infectious disease in their previous clinical practice. Although it is reasonable that training and education may benefit to reduce negative effects caused by goggles and have been emphasized in previous studies^[11]^, however, our findings showed short-term education and training on wearing PPE has limited effectiveness on easing mental stress and discomforts and cannot substantially avoid the occurrence of medical errors. A large number of nurses in all groups occurred goggle-associated discomforts and had experiences of discontinuation from work at some time.

Although goggle-associated symptoms widely presented in all of nurses, we found nurses with≥40 years or≥21 years working experience showed a more optimistic attitude to the impact of mental stress, discomforts or fog on working status and the occurrence of medical errors. In addition, the proportions of headache and vomit were also significant lower in nurses with ≥21 years working experience. Because both of two groups represent more mature mentality and more abundant working experience, furthermore, age and practice duration were highly correlated with each other in our study (r = 0.94, p < 0.001, Supplement Fig 2). The findings revealed that training on psychological enduring capacity and enhancing proficiency may be useful to overcome some symptoms that can be induced by stress and can be more quickly adapting to new situations in a shorter time. Considering training is the only way to increase the experience in short time, though short-term training was found to have limited effects on improving mental stress, we strongly suggest longer and earlier training and education should be performed to help the accumulation of experience with working in PPE more rapidly, especially in younger nurses.

Headache was mostly mentioned in PPE-associated symptoms in previous study^[10]^. Our results supported that headache could be the most prevalent symptom caused by goggles. Moreover, dizziness, nausea, skin pressure injury, vomit and dysphoria were also broadly present in nurses during practice in management of COVID-19. Interestingly, unlike the distribution of most symptoms in this study, the incidence of rash was much higher in nurses with≥21 years working experience, the possible reason is the skin was aging in this group and was more susceptive to the hostile environment, indicating the prevention of symptoms should be performed discretely on nurses in different age range.

Goggle-associated discomforts occurred in early stage and these discomforts seriously impacted their clinical practice during management of patients with COVID-19 and contributed to medical errors. Moreover, it is worthy to notice that most of nurses had to adjust their goggles during work even they knew this action would increase the exposure risk. Our data demonstrated that discomforts are not only harmful to the health but also lead to inferior quality of clinical works, thus relief of these discomforts is essential to protect healthcare professionals and patients. For the underlying reasons to cause these discomforts, tightness of goggles was complained mostly. Wearing too tight can be impacted by subjective reasons and objective reasons. As we described previously, mental stress on fearing exposure partly made nurses to wear goggles more tight than ordinary time. On the other hand, some nurses also attenuated discomforts through selecting softer band or wider band and using anti-stress trips to reduce the pressure, indicating materials and designs may also be useful to improve the user experience on tightness. In addition, besides to tightness feeling, we also identified the unsuitable face shape and hard materials used for producing goggles were also two important reasons to induce the discomforts. Consistent with previous conclusion, adequate psychological training is essential to help resolving mental stress-associated discomforts. Moreover, the design and materials can be substantially improved, a customized goggle for different races with softer band or wider band may be more popular in nurses.

Medical errors can potentially cause serious problems during clinical practice. Unfortunately, our study found goggle is associated with increased medical errors and this cannot be changed significantly by more abundant working experience. Among the risk factors, the longer time before tolerating discomforts and headache meant the nurses’ focus on practice would be interrupted and more frequent adjusting goggles can increase the chances of occurring medical errors. Because these three risk factors all associated with mental stress and design of goggles, modified goggles and psychological education should be seriously considered.

Fog is another refractory problem of goggles. In this study, the median time to arising fog in goggles was only 1 hour after wearing PPE, indicating nurses had to work with blurred vision for a long time. Alarmingly, our results revealed high proportion of nurses considered fog in goggles substantially impacted their work and associated with medical errors. Although we did not compare the incidence of exposures caused by fog in management of COVID-19 with the incidence in ordinary times, the exposure incidence of 10.4% cannot be neglected. Although several methods had been proven in clinical practice, especially smearing some types of reagents. The rating on the efficiency varied in groups, we noticed that the efficient of measures taken by younger nurses was inferior, the reason may be that younger nurses had less skilled. In terms of these discrepancy of efficiency, we suggested if similar measures can be considered and adopted during productive process, the effects would be more reliable.

There are some limits in this study: First, the sample size was a bit small. However, all of age groups and working experience were covered in this study and our results can reflect the overall condition. Second, this survey was performed after mission completed and the participants have rested for several days, this may cause potential bias on rating. The negative effects and the level of nervous may be underestimated. Third, this retrospective study performs a preliminary assessment, goggle-associated symptoms and negative effects of fog in goggles were primary analyzed, the effects of other PPE on health and working status of healthcare professionals should be further investigated.

Taking together, goggle-associated symptoms and fog are two major problems that impact the clinical practice. Goggle-associated symptoms are highly prevalent in nurses involving in management of patients with COVID-19. These discomforts significantly impacted working status and potentially lead to medical errors. Headache, adjusting goggles and time before adapting goggles are three risk factors of medical errors. On the other hand, fog can obviously interfere the vision and lead to medical exposures. Improvement of design and materials of goggles during productive process is promising way to reduce the incidence of symptoms and fog and may benefit to working status and the occurrence of medical errors. In addition, longer training time and psychological education may help to reduce the mental stress through increasing the psychological quality and proficiency.

## Data Availability

The authors confirm that the data supporting the findings of this study are available within the article and its supplementary materials.

## Authors’ contributions

HXH, FYR, PXJ, CT, ZYL, ZH, LJ, LLM, FL, HX, WYH performed surveys and were responsible for data collection. KX, WYH, LGM, HXH, FYR, PXJ designed the questionnaires. KX, LGM prepared data. LGM, KX, MHM, HXH performed statistical analysis and data interpretation. HXH, KX, MHM, FYR, LGM, PXJ wrote manuscript. KX, MHM, WYH, Zheng Wei conceived the study, designed the survey process. All authors read and approved the final manuscript.

## Acknowledgements

The authors would like to thank all healthcare professionals for the efforts in fighting against SARS-CoV2 and all scientists who work for developing treatment of COVID-19 and who share their research works about this pandemic.

